# Study protocol of the Hungarian Longitudinal Study of Healthy Brain Aging (HuBA)

**DOI:** 10.1101/2023.11.09.23298159

**Authors:** Éva M. Bankó, Béla Weiss, István Hevesi, Annamária Manga, Pál Vakli, Menta Havadi-Nagy, Rebeka Kelemen, Eszter Somogyi, István Homolya, Adél Bihari, Ádám Simon, Ádám Nárai, Krisztina Tóth, Noémi Báthori, Vivien Tomacsek, Anita Kamondi, Mihály Racsmány, Ádám Dénes, Péter Simor, Tibor Kovács, Petra Hermann, Zoltán Vidnyánszky

**Author notes:** Corresponding authors: Éva M. Bankó, Brain Imaging Centre, HUN-REN Research Centre for Natural Sciences, Magyar tudósok krt. 2, 1117, Budapest, Hungary, Zoltán Vidnyánszky, Brain Imaging Centre, HUN-REN Research Centre for Natural Sciences, Magyar tudósok krt. 2, 1117, Budapest, Hungary.

## Abstract

**Background:** Neurocognitive aging and the associated brain diseases impose a major social and economic burden. Therefore, substantial efforts have been put into revealing the lifestyle, neurobiological and genetic underpinnings of healthy neurocognitive aging. However, these studies take place almost exclusively in a limited number of highly-developed countries. Thus, it is an important open question to what extent their findings may generalize to neurocognitive aging in other, not yet investigated regions.

**Purpose:** The purpose of the Hungarian Longitudinal Study of Healthy Brain Aging (HuBA) is to collect multi-modal longitudinal data on healthy neurocognitive aging to address the data gap in this field in Central and Eastern Europe.

**Methods:** We adapted the Australian Imaging, Biomarkers and Lifestyle (AIBL) study of aging^1^ study protocol to local circumstances and will collect demographic, lifestyle, mental and physical health, medication and medical history related information as well as record a series of magnetic resonance imaging (MRI) data. In addition, participants will also be offered to participate in the collection of blood samples to assess circulating inflammatory biomarkers as well as a sleep study aimed at evaluating the general sleep quality based on multi-day collection of subjective sleep questionnaires and whole-night electroencephalographic (EEG) data.

**Results & Discussion:** Data collection will be longitudinal with 18 months between measurements and at least three sessions are intended. The collected data might reveal specific local trends or could also indicate the generalizability of previous findings. Moreover, as the HuBA protocol also offers a sleep study designed for thorough characterization of participants’ sleep quality and related factors, our extended multi-modal dataset might provide a base for incorporating these measures into healthy and clinical aging research.

**Conclusion:** Besides its straightforward national benefits in terms of health expenditure, we hope that this Hungarian initiative could provide results valid for the whole Central and Eastern European region and could also promote aging and Alzheimer’s disease research in these countries.

## Introduction

Stable economic background supports the development of social and health services as well as nutrition that in turn contribute to increase of life expectancy and number of years spent in health. As a result, during the last century, life expectancy has increased by several decades in some countries ^2^. However, increase of life expectancy and certain lifestyle changes increases also the prevalence of various neurocognitive disabilities such as Alzheimer’s disease (AD), which, together with the demographic shift from young to old, is placing a significant socio-economic burden on the societies concerned.

To address these issues, major efforts are underway to foster research on healthy aging and understanding the lifestyle correlates, genetic underpinnings and neurobiological mechanisms of AD, its prevention, diagnosis, and treatment related to this devastating disease ^1–7^. Although the aforementioned trends in life expectancy, demographic shift and prevalence of AD are most prevalent in developed countries, these tendencies are present also in countries all around the globe. However, the large-scale research programs on healthy aging and AD are exclusively run in highly developed countries making thus the sampling not representative globally. As these studies apply complex investigation approaches relying on multi-modal data including also detailed demographic and lifestyle measures, the generalizability of their findings to unrepresented world regions is an open question due to significant alterations in these variables stemming from significantly different economic, cultural and historical backgrounds.

Here we introduce the Hungarian Longitudinal Study of Healthy Brain Aging (HuBA) that is aimed at addressing the backlog in the field of healthy aging in Hungary. The primary goal of this study is to adapt the complex investigation approaches used by the abovementioned research programs in developed countries as well as to collect an initial multi-modal cohort dataset that could be used for testing of data harmonization methods ^8,9^ and could also serve the optimization of study protocols before scaling up data collection. As a reference for study protocol design, the Australian Imaging, Biomarkers and Lifestyle (AIBL) study of aging^1^ was considered. After adapting the AIBL procedures to local circumstances, we ended up with a study protocol comprising a collection of demographic, lifestyle, mental and physical health, medication and medical history related information as well as recording a series of magnetic resonance imaging (MRI) data. In addition to these compulsory data modalities, participants will also be offered to participate in the collection of blood samples as well as a sleep study aimed at assessing the general sleep quality based on multi-day collection of subjective sleep questionnaires and whole-night electroencephalographic (EEG) data. As most data harmonization methods require site-level estimation of central tendencies for elimination of batch effects ^8,9^ to allow for harmonizing the recorded data with those collected in other studies, the initial sample size was set to N=100 participants, and the time interval between the baseline and first follow-up measurements was chosen to be 18 months in agreement with other longitudinal studies.

Closely following the international data collection and analysis standards developed for healthy aging and AD cohort studies, we believe that after harmonization, our data could contribute to the field by indicating local trends if such exist, and would also pave the way for national aging research. Besides its clear national benefits, having a similar economic, cultural and historical background with other countries in Central and Eastern Europe, we hope that this Hungarian initiative could provide results valid for the whole region and could also promote aging and AD research in these countries.

## Methods

### PARTICIPANTS

#### 1. Recruitment and selection

Participants will be recruited with the help of a short study description from the local community through mailing lists and Facebook posts. The planned sample size is 100 participants. All potential participants will be screened through a phone interview, where they will be informed of the goals and details of the study, and will be screened for eligibility. Participants have to be cognitively healthy, not have a serious psychiatric condition, and be eligible for MRI scanning. They also have to indicate their willingness to participate in all measurements, except for blood samples and sleep research, which are not definitive requirements for eligibility. Detailed eligibility criteria can be found in Table 1. All participants will be required to provide an informed consent and they will also be informed that they will be free to withdraw at any stage after starting the study. The study flowchart can be seen in Figure 1. This study will be conducted in compliance with the Helsinki Declaration, and has been approved by the Hungarian National Institute of Pharmacy and Nutrition (reference#: OGYÉI/68903/2020), which has the authority of overseeing medical research in Hungary.

**Table 1.**
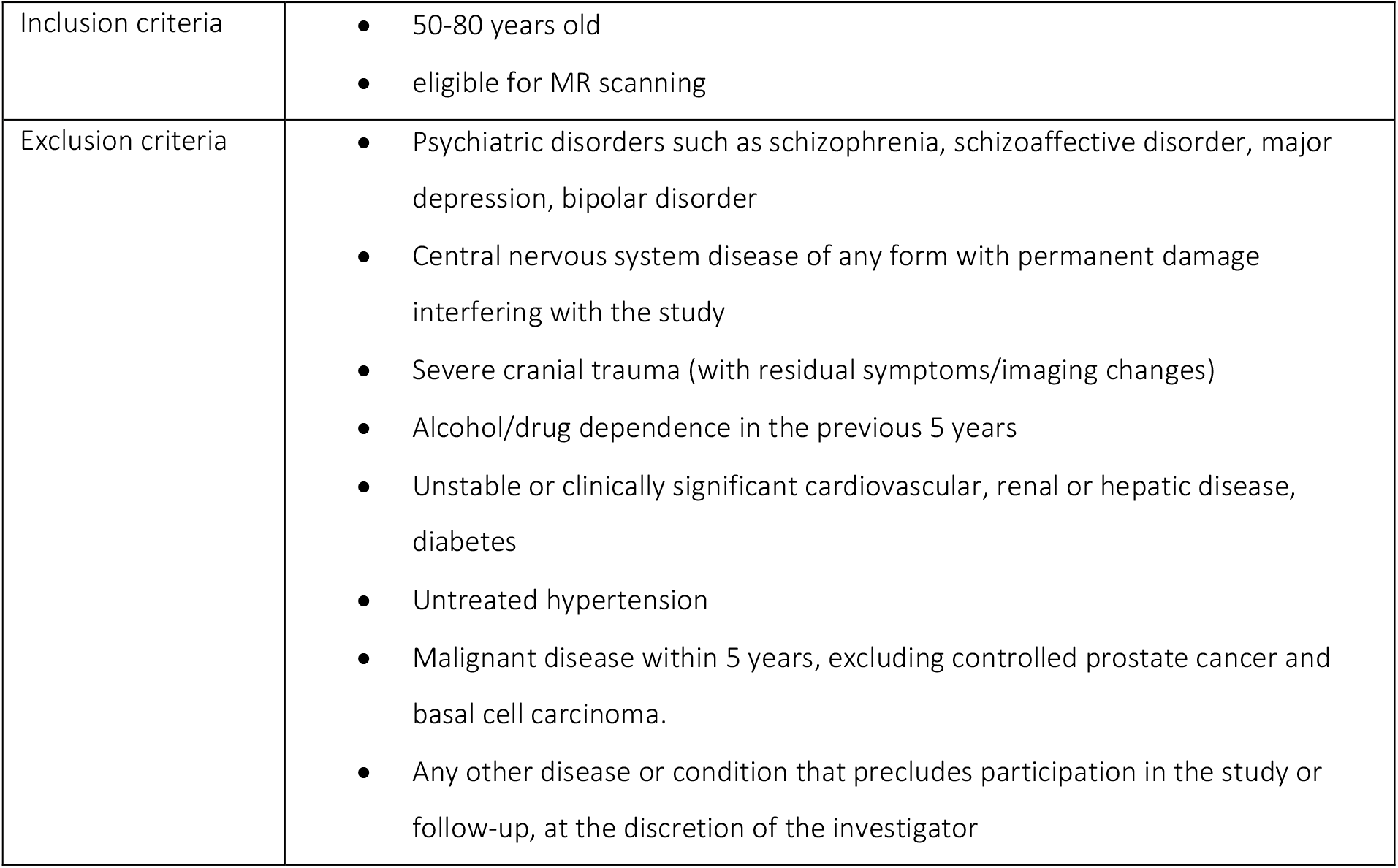
Inclusion and exclusion criteria.

**Figure 1.**
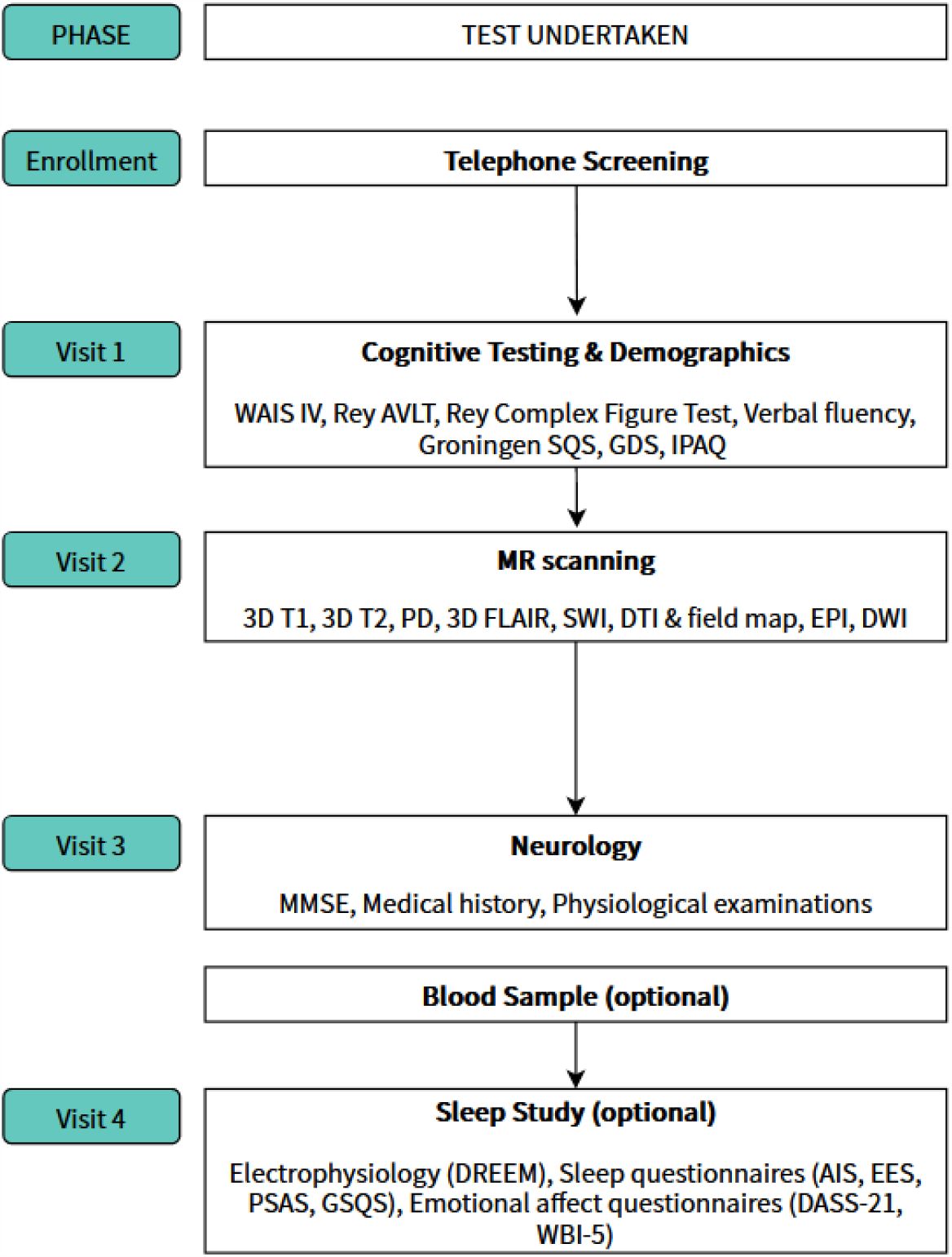
Flowchart of study design with participant recruitment and participation at each stage

### VISIT 1: COGNITIVE TESTING

#### 1. Demographic information

Basic participant information will be recorded on the baseline session only, including date of birth, gender, highest education and years spent in formal education, employment and profession. In addition, handedness, as assessed by the Edinburgh Handedness Inventory ^10^ will also be recorded.

#### 2. Baseline IQ

Participants will be required to complete the fourth edition of the Wechsler Adult Intelligence Scale (WAIS-IV; Giunti Psychometrics Hungary, Budapest, Hungary) for obtaining a total IQ measure at the beginning of the study, which will not be repeated on follow-up sessions. Instead, from Session 2 onward only the Digit Span and Digit Symbol Coding subtests will be administered (see below).

#### 3. Cognitive and mood assessments

After a short introduction and briefing on the technical details of the testing session, participants will be examined using a battery of tests. First, they complete the Digit Span and Digit Symbol Coding subtests of the fourth edition of the Wechsler Adult Intelligence Scale (WAIS-IV; Giunti Psychometrics Hungary, Budapest, Hungary). Next, the immediate recall and interference trials of the Rey Auditory Verbal Learning Test (AVLT) ^11,12^ will be completed, followed by the Rey Complex Figure Test’s (RCFT) ^13^ copy and immediate recall tasks. During the delay periods of both the AVLT and RCFT, participants fill out the Groningen Sleep Quality Scale (GSQS) ^14,15^, Geriatric Depression Scale (GDS) ^16,17^, all of which are administered in a standard pen and paper format. If the delay periods of the previous tests are still yet to expire, participants will be let on a short break, but care will be taken that they not engage in any mentally demanding tasks in the remaining time, in order to prevent interference with the tests. After the delay, the session resumes with the delayed recall task of the RCFT, then with the delayed recall trial of the AVLT. Finally, participants will be administered a verbal fluency task (category fluency: animals, letter fluency: ‘m’).

#### 4. Life style & physical health

Participants also fill out a questionnaire relating to lifestyle in the first measurement session only. They provide information about physical wellbeing, social contact and on aspects of their diets. To get a proxy for weekly physical activity, participants also complete the short version of the International Physical Activity Questionnaire ^18,19^ and total metabolic equivalent (MET) minutes per week will be calculated. In addition, participants answer questions related to subjective sleep time as in items 1,3, and 4 of the Pittsburgh Sleep Quality Index (PSQI, ^20,21^ and subjective sleep efficiency will be calculated as time spent with sleep divided by time spent in bed.

### VISIT 2: MRI SESSION

Collection of MRI data will be performed at a single site, the Brain Imaging Centre, Research Centre for Natural Sciences, Budapest. MR measures are gathered in a one-hour session conducted on a 3T Siemens Prisma system, employing a 32 channel head coil.

Measures in the MRI session included:

#### 3D T1-weighted structural image

A high resolution 3D T1-weighted structural image is acquired using a Magnetization Prepared RApid Gradient Echo (MPRAGE) sequence with the following parameters: Repetition Time (TR) =2300 milliseconds; Echo Time (TE) =3.03 milliseconds; Inversion Time (TI) =900 milliseconds; flip angle =9 degrees; field of view (FOV) =256mm x 256mm x 192mm; voxel size =1mm isotropic; GeneRalized Autocalibrating Partial Parallel Acquisition (GRAPPA) acceleration factor =2; acquisition time of 5 minutes and 21 seconds.

#### 3D T2-weighted structural image

A high-resolution 3D T2-weighted structural image is acquired with a SPACE sequence with the following parameters: TR =3200 milliseconds; TE =558 milliseconds; FOV =240mm × 256mm × 176mm; resolution =1mm isotropic; GRAPPA acceleration factor =2; acquisition time of 5 minutes and 47 seconds.

#### PD-weighted structural image

A high resolution 2D coronal proton density weighted turbo spin echo image is acquired from the hippocampal regions (slices are perpendicular to the right hippocampus) with the following parameters: TR =6490 milliseconds; TE =16 milliseconds; FOV =206mm × 165mm × 66mm; resolution =0.4 × 0.4 × 2mm ; Flip Angle: 120 degree; Distance Factor: 0% ; Turbo factor:11; Echo Spacing: 15.7ms, Echo Trains per Slice: 38; acquisition time of 4 minutes and 15 seconds.

#### 3D FLAIR structural image

A high-resolution 3D FLAIR (Fluid Attenuated Inversion Recovery) structural image is acquired with a SPACE sequence with the following parameters: TR =5000 milliseconds; TE =386 milliseconds; FOV =240mm × 256mm × 176mm; resolution =1mm isotropic; GRAPPA acceleration factor =2; acquisition time of 5 minutes and 37 seconds.

#### Susceptibility Weighted Image (SWI)

Axial Susceptibility Weighted Images are acquired with the following parameters: TR =27 milliseconds; TE =20 milliseconds; FOV =220mm × 200mm × 144mm; resolution =0.9 x0.9 × 1.5mm; Flip Angle: 15 degree; Distance Factor: 20%; GRAPPA acceleration factor =2; acquisition time of 5 minutes and 24 seconds.

#### Field map for the DTI

We acquired an axial field map with the following parameters: TR =400 milliseconds; TE =4.92 and 7.38 milliseconds; FOV =208mm × 208mm × 135mm; resolution =3mm isotropic; Flip Angle: 60 degree; Distance Factor: 25%; Acquisition time of 58 sec. Diffusion-Weighted Images (DWI).

#### Diffusion Tensor Imaging (DTI)

Axial DTI images are acquired with the following parameters: b-values: 0 s/mm^2^, 2000 s/mm^2^ with 109 gradient directions; TR =2800 milliseconds; TE =68 milliseconds; FOV =208mm × 208mm × 120 mm; resolution =2mm isotropic; Distance Factor: 0%; SMS Acceleration Factor:4; Phase Partial Fourier:6/8; acquisition time of 5 minutes and 24 seconds.

#### Segmented EPI

Axial segmented EPI image is acquired from the midbrain region with the following parameters: TR =150 milliseconds; TE =36 milliseconds; FOV =180mm × 240mm × 192mm; resolution =0.4 × 0.4 × 1mm; Distance Factor: 50%; Flip Angle: 30 degree; acquisition time of 2 minutes and 08 seconds.

#### Diffusion-Weighted Image

Axial diffusion-weighted Images (DWIs) are acquired with an EPI sequence, with 3 b-values: 0 s/mm^2^, 500 s/mm^2^ and 1000 s/mm^2^ (with 2, 3, 3 averages, respectively). TR =4800 milliseconds, TE =79 milliseconds, voxel size =0.4 × 0.4 × 4 mm, FOV =220 mm × 200 mm x 160mm; Distance factor 30% ; acquisition time of 1 minutes and 50 seconds

### VISIT 3: NEUROLOGY

Visit 3 is going to be conducted at the Department of Neurology, Semmelweis University, Budapest, Hungary.

#### 1. Mental health, medical history and medication use

All participants undergo a complete neurological examination where their full neurologic status will be evaluated. In addition, information on personal medical history, medication use, smoking, current and past alcohol use is collected along with family history of dementia and other neurological illnesses. The Mini Mental State Examination (MMSE) (Folstein et al., 1975) is conducted to assess sufficient mental health to be part of the cohort.

#### 2. Physiological measures

In order to assess core physiological factors that affect cardiovascular health, participants’ height and weight are also measured along with their pulse and blood pressure. In addition, participants’ internal medical status is also recorded. All measurements will be done by the evaluating neurologist.

#### 3. Blood samples

From participants who consent, blood samples will be also taken at this stage. Peripheral venal blood is collected to K-EDTA tubes. Part of it will be forwarded to a clinical laboratory (Semmelweis Egyészségügyi Kft., Budapest, Hungary) for apolipoprotein E genotyping, B12 and TSH measurements. Sample processing is performed as described previously ^23^. The rest of the anticoagulated blood is fractioned into plasma, platelets, red blood cells, and white blood cells by centrifuging at 2000g at 4°C for 10 minutes within 30 minutes of obtaining. The plasma will subsequently be stored at -80°C for further biomarker analysis. To examine the associations of circulating inflammatory biomarkers with neurocognitive aging, the concentration of the key inflammatory cytokines and chemokines (TNFα, IFNγ, IL-17A, IL-6, IL-1α, IL-1β, IL-8, ICAM-1, P-selectin, CD121a, CD121b, MCP-1, RANTES, Angiogenin and Fractalkine) is determined by BD Cytometric Bead Array (CBA) in the blood plasma using BD CBA Flex Sets according to the manufacturer’s instructions.

### VISIT 4: SLEEP STUDY

Visit 4 takes place either in the Brain Imaging Centre (Research Centre for Natural Sciences, Budapest, Hungary) or in the Budapest Laboratory of Sleep and Cognition (Eötvös Loránd University, Budapest, Hungary).

#### Pre-visit

Participants fill in a cross-sectional test battery online, which containes the following questions and questionnaires:

- Athens Insomnia Scale (AIS) ^24^
- Epworth Sleepiness Scale (ESS) ^25^
- Pre-Sleep Arousal Scale (PSAS) ^26^
- Depression, Anxiety and Stress Scale (DASS-21) ^27^
- questions about post-COVID symptoms rated on a 10-point Likert scale (1 = Not at all; 10 = Very much) (autonomic, musculoskeletal, neurological, cognitive, psychological, gastrointestinal symptoms, olfactory and/or gustatory disturbances, sleep disturbances);
- 8 questions about perceived cognitive failures rated on a 5-point Likert scale (0 = Never; 4 = Very often) based on the Cognitive Failures Questionnaire (CFQ) ^28^;
- WHO Well-Being Questionnaire (WBI-5) ^29^

#### Onsite

Participants are educated on the protocol of the seven-day-long home-based sleep study. The study, and hence the training consists of two parts: (1) assessment of subjective experiences throughout the day, and (2) physiological measurement at night (see the Sleep study subsection for details). An information sheet with a checklist aids them in all the steps and tasks over the course of a day regarding both parts.

##### 1. Assessment of subjective experiences throughout the day

Participants are already familiar with the online platform where the questionnaires could be accessed. They are informed when they get notified of each questionnaire via email and when they should fill them in.

##### 2. Physiological measurement at night

Participants are trained on the use of the device, that is, a portable EEG headband (Dreem 2 Headband, Rhythm, Paris, France) and its smartphone application named Alfin. After downloading the application on their own smartphone and signing in with their own account, they are asked to put on the device, helped how to position it properly and shown how to start a recording via the application (i.e., evening protocol). After a sample recording, they are explained how to stop a recording, upload it and charge the device (i.e., morning protocol). A detailed paper-based manual and several videos provides them with further possible offsite assistance on all the steps. To enhance signal quality, they are also given a sweatband to keep the headband closer to the skin and the scalp, and isopropyl alcohol wipes to clean their skin and the sensors daily.

#### Sleep study

The prospective sleep study lastes for seven days; however, it startes on day 0 with an afternoon questionnaire and endes on day 7 also with an afternoon questionnaire. During the seven days, participants report on their subjective experiences three times a day and wear a portable EEG headband at night. The notifications for the questionnaires are sent at specific times via emails (the morning questionnaire at 5:30 a.m., the afternoon questionnaire at 5:00 p.m., and the evening questionnaire at 9:00 p.m.), but participants are asked to fill them in upon awakening, between 5:00 and 9:00 p.m. (at least two hours before the evening questionnaire), and before going to bed, respectively. Responses with a certain extent of deviation will be excluded from the analyses, that is, more than two hours after the end of the EEG recording in case of the morning questionnaire; after 11 p.m. in case of the afternoon questionnaire; and not right before the beginning of the EEG recording in case of the evening questionnaire. Timestamps indicating the beginning and the end of an EEG recording, as well as the exact time when a questionnaire was filled in will be derived from the output files available on the servers of the headband and questionnaires.

The prospective assessments are carried out with the following instruments:

##### 1. The assessment of subjective experiences

a. <u>Morning questionnaire</u>:
  - question(s) about dreaming; in case of dream recall, additional items: dream content, the rating of 13 dream emotions on a 5-point Likert scale (1 = Not at all; 5 = Very much) (joy, guilt, satisfaction, surprise, fear, curiosity, sadness, safety, anger, shame, insecurity, disgust, excitement), nightmare occurrence
  - questions about sleep: nodding off, time of going to bed, subjective sleep onset latency, time of waking up;
  - Groningen Sleep Quality Scale ^14^, modified: ratings on a 5-point Likert scale (0 = Strongly disagree; 4 = Strongly agree) instead of binary response options;
  - a question assessing vigilance on an 8-point Likert scale (1 = I am extremely vigilant; 8 = I am very sleepy, I cannot stay awake);
  - the rating of 12 emotions on a 7-point Likert scale (1 = Not at all; 7 = Very much): (joy, excitement, sadness, satisfaction, fear, worry, activity and vigour, irritability, displeasure, restlessness, confusion, anger);
  - a question about taking sedatives or sleeping pills/drugs/herbs;
  - a question about the name and the amount of the specific drug(s).
b. <u>Afternoon questionnaire</u>:
  - Psychotic-Like Experiences ^30^;
  - a question about the length of potential napping (in minutes) (in case of napping they do not have to wear the headband);
  - the rating of 12 emotions on a 7-point Likert scale (1 = Not at all; 7 = Very much): (joy, excitement, sadness, satisfaction, fear, worry, activity and vigour, irritability, displeasure, restlessness, confusion, anger);
  - questions about post-COVID symptoms rated on a 10-point Likert scale (1 = Not at all; 10 = Very much) (autonomic, musculoskeletal, neurological, cognitive, psychological, gastrointestinal symptoms, olfactory and/or gustatory disturbances, sleep disturbances);
  - 8 questions about perceived cognitive errors rated on a 5-point Likert scale (0 = Never; 4 = Very often).
c. <u>Evening questionnaire</u>:
  - Pre-Sleep Arousal Scale (PSAS) ^26^;
  - questions about alcohol and caffeine intake.

##### 2. Physiological measurement at night

The mobile EEG headband can be operated with its phone application, and uploads data to the server via Wi-Fi. The headband records five types of data using three types of sensors: (1) cortical activity (250 Hz; filtered between 0.4 and 35 Hz) via five dry EEG electrodes (O1, O2, FpZ, F7, F8); (2-4) movement, body position and respiratory rhythm using a 3D accelerometer placed above the head; (5) pulse, detected by an infrared pulse oximeter placed on the frontal strap. The EEG electrodes are made of solid silicone and have soft, flexible protrusions on the occipital lobe to capture the signal through the hair. The headband is made of foam and fabric and has an adjustable strap to make it comfortable to use and to keep the electrodes close enough to the scalp ^31^.

The headband also offers a machine-learning based, automated sleep staging that demonstrated equivalent performance to human raters scoring the polysomnographic recordings in a study involving the simultaneous assessment of polysomnographic recordings and recordings carried out with the Dreem headband ^31^. Therefore, it is also possible to download the hypnogram of each night after uploading a file.

The protocol of the sleep study in summary is the following: when participants wake up, they end the EEG recording, start to charge the device, and while the data are being uploaded, they complet the morning questionnaire. In the afternoon, they answer the questions of the afternoon questionnaire. In the evening, when they decide on going to bed, they clean their skin and the sensors of the headband with a wipe, put it on their head, cover it with a sweatband, send their responses of the evening questionnaire, and start the EEG recording with their smartphone.

#### Offsite

Since participants will upload the recordings each morning, it will be possible to verify after each night if the signal quality was appropriate. In case of lower quality, participants will be contacted via emails or phone calls and instructed how to improve the placement of the headband and thus to enhance the quality of the recordings. No other information about their recordings will be disclosed to the participants during data collection.

#### Post-sleep study visit

Upon completion of the seven-day-long study, participants bring back the device and are given the hypnograms of their nights generated by the algorithm. Participants have the opportunity to share their experiences about the sleep study, give feedback and ask for contact details of a sleep clinic if necessary.

## Discussion

Life expectancy has significantly increased over the past century. As a result, the prevalence of Alzheimer’s disease (AD) has also dramatically increased, and its social, economic and health care impact has become enormous. Therefore, substantial efforts have been put into revealing the lifestyle, genetic and neurobiological underpinnings of healthy neurocognitive aging and AD. These research programs have already contributed a lot to the understanding of aging and AD disease processes by revealing new markers that could extend further the currently available diagnostic and therapeutic approaches. However, these studies take place almost exclusively in a limited number of highly-developed countries. Thus, it is an important open question to what extent their findings may generalize to neurocognitive aging in other, not yet investigated regions. The aim of the HuBA study introduced in this paper is to address this limitation by implementing the currently available international standards of healthy aging research in Hungary. To this end, the protocol of the AIBL study was adapted and data collection for at least a hundred participants is planned. The collected data, after proper harmonization with data from previous studies, might reveal specific local trends and also indicate the generalizability of previous findings. Moreover, as the HuBA protocol also offers a sleep study designed for thorough characterization of participants’ sleep quality and related factors, our extended multi-modal dataset might provide a base for incorporating these measures into healthy and clinical aging research. Besides its straightforward national benefits, we hope that our study can also promote and advance aging research in the broader Central European region.

## Data Availability

No data is presented in this protocol manuscript

## Competing interests

The authors declare that they have no competing interests.

## Acknowledgements

This research was supported by the Hungarian National Research, Development and Innovation Office (grant number 2019-2.1.7-ERA-NET-2020-00008) and Project no. RRF-2.3.1-21-2022-00015, which has been implemented with the support provided by the European Union. This work was also supported by a grant from HUN-REN (0708-21 515 AT) to MR and ZV. We thank Szandra Pancsusák and Emília Nagy for managing patient logistics and are very grateful to all participants for their time and participation in this study.

## Notes

### Competing Interest Statement

The authors have declared no competing interest.

### Author Declarations

This study will be conducted in compliance with the Helsinki Declaration, and has been approved by the Hungarian National Institute of Pharmacy and Nutrition (reference#: OGYÉI/68903/2020), which has the authority of overseeing medical research in Hungary.

